# Effects of hydrometeorological and other factors on SARS-CoV-2 reproduction number in three contiguous countries of Tropical Andean South America: a spatiotemporally disaggregated time series analysis

**DOI:** 10.1101/2022.06.13.22276339

**Authors:** Josh M. Colston, Patrick Hinson, Nhat-Lan H. Nguyen, Yen Ting Chen, Hamada S. Badr, Gaige H. Kerr, Lauren M. Gardner, David N. Martin, Antonio M. Quispe, Francesca Schiaffino, Margaret N. Kosek, Benjamin F. Zaitchik

## Abstract

**Background:** The COVID-19 pandemic has caused societal disruption globally and South America has been hit harder than other lower-income regions. This study modeled effects of 6 weather variables on district-level SARS-CoV-2 reproduction numbers (R_*t*_) in three contiguous countries of Tropical Andean South America (Colombia, Ecuador, and Peru), adjusting for environmental, policy, healthcare infrastructural and other factors.

**Methods:** Daily time-series data on SARS-CoV-2 infections were sourced from health authorities of the three countries at the smallest available administrative level. R_*t*_ values were calculated and merged by date and unit ID with variables from a Unified COVID-19 dataset and other publicly available sources for May – December 2020. Generalized additive mixed effects models were fitted.

**Findings:** Relative humidity and solar radiation were inversely associated with SARS-CoV-2 R_*t*_. Days with radiation above 1,000 KJ/m^2^ saw a 1.3%, and those with humidity above 50%, a 1.0% reduction in R_*t*_. Transmission was highest in densely populated districts, and lowest in districts with poor healthcare access and on days with least population mobility. Temperature, region, aggregate government policy response and population age structure had little impact. The fully adjusted model explained 3.9% of R_*t*_ variance.

**Interpretation:** Dry atmospheric conditions of low humidity increase, and higher solar radiation decrease district-level SARS-CoV-2 reproduction numbers, effects that are comparable in magnitude to population factors like lockdown compliance. Weather monitoring could be incorporated into disease surveillance and early warning systems in conjunction with more established risk indicators and surveillance measures.

**Funding:** NASA’s Group on Earth Observations Work Programme (16-GEO16-0047).

## Introduction

Since its discovery in Wuhan, China in December 2019, the SARS-CoV-2 virus has swept the globe, overwhelming national healthcare services in successive waves and variants, and causing widespread socio-economic insecurity and societal unrest in virtually every country of the world.^1,2^ As of the time of writing, over 533 million infections and 6.3 million deaths globally, have been attributed to the virus^3^, though the true toll is undoubtedly far higher than official statistics, and may have surpassed 3.8 billion infections (40% of the global population) and 15 million deaths.^4^ South America has been hit harder by the Coronavirus disease (COVID-19) pandemic than other predominantly lower income regions with some of the highest excess mortality and case fatality rates (CFR), its 58 million confirmed cases (>256 million estimated total) leading to over 1.3 million confirmed deaths (>1.7 million total), putting further strain on a region where many countries struggle with political instability, humanitarian crises, and income inequality.^3–6^ From the early days of the pandemic, questions were raised about the possible influences of climate and meteorology on the transmission of the virus given the known sensitivity of other respiratory viruses to these factors.^7,8^ One early study noted that COVID-19 community transmission at the beginning of the pandemic was especially high along a temperate mid-latitude belt of the northern hemisphere.^9^ However, it was already clear by that early stage that the influence of such factors was small relative to that of population density and age structure, and timing of and compliance with non-pharmaceutical interventions (NPIs) such as lockdowns, travel restrictions and hygiene measures, and initial research rightly prioritized these more proximal drivers.^10–12^

With the pandemic in its third year, and with the likely prospect that SARS-CoV-2 will continue to circulate as an endemic, seasonal and vaccine-preventable virus for the foreseeable future^13^, attention has turned again to the role of meteorological factors in COVID-19 transmission.^8,14^ The demand for real-time data with which to track the global health crisis has prompted a proliferation of online repositories and interfaces, which curate and disseminate epidemiological data with global scope and increasing spatial and temporal resolutions.^6,15,16^ Disease data can be matched by date and location to high resolution estimates of spatiotemporal variation in environmental and hydrological conditions derived from remote sensing and climate models for further analysis.^17^ Numerous studies have applied this approach to subnational unit-level case reports in an attempt to model associations between hydrometeorological variables and SARS-CoV-2 outcomes^18,19^, however there is considerable variation in how confounding factors and error in case reporting are captured^20,21^, and a disproportionate emphasis on High Income Countries, mostly in the temperate mid-latitudes.^22^

The aim of this study was to model the effects of weather on the district-level SARS-CoV-2 reproduction number (R_*t*_) for three contiguous countries of Tropical Andean South America (Colombia, Ecuador, and Peru), with an expanded suite of hydrometeorological parameters and after further adjusting for environmental, policy, healthcare infrastructural and other factors during the first wave of the epidemic, when a single circulating variant predominated and there was no population level immunity that contributed to transmission dynamics..

## Methods

### Scope of Analysis

The three Tropical Andean South American countries of Colombia, Ecuador and Peru were chosen for this analysis, since together they constitute a large contiguous territory, roughly evenly split between the northern and southern hemispheres and broadly divisible into coastal, highland, and interior regions. Furthermore, all three countries have comparable health information system capacity and make publicly available daily reports of new COVID-19 cases at highly geographically disaggregated levels. The analysis was restricted to the mainland areas of the three countries, excluding outlying island territories, and to the period from May to December 2020, during which transmission of the virus was fully established and NPIs in place^19^, but before the emergence of major variants of concern and the introduction of vaccines.

### Epidemiological Data

Daily time series data on confirmed SARS-CoV-2 infections were sourced from national health authority websites at the smallest available administrative level (Colombian municipalities, Ecuadorian cantons and Peruvian districts, hereafter generically referred to as “districts”).^23–25^ These data were used to calculate district-level daily R_*t*_ using EpiNow2, an R package for estimating time-varying epidemiological parameters of SARS-CoV-2 from subnational case notification data accounting for right truncation, underreporting and uncertain reporting delays and incubation periods.^26^ Daily, district-level R_*t*_ estimates were treated as the outcome variable for the analysis and are interpreted as the mean number of new infections caused by a single infected person on a given day in a given district. If a district records zero cases for an extended period, its daily R_*t*_ will converge on a default value of 1, which is difficult to interpret in the absence of actual disease. However, because the calculation of R_*t*_ accounts for the disease incubation period, the metric lags the cases used to calculate it, so changes in R_*t*_ may precede increases and decreases in case counts by several weeks. It is therefore possible for a district to have a daily R_*t*_ of greater than 1 while reporting zero cases, due to the delay in increases in transmission being reflected in case reporting. Due to the high resolution and inclusion of many remote and sparsely populated districts in the dataset, there was a large proportion of unit-days with zero reported cases of COVID-19 (75.5%). We therefore excluded all unit-days in which both: a). no cases were reported and b). R_*t*_ had a calculated value of between 0.95 and 1.05, with the purpose of restricting the analysis to observations with interpretable outcome values.

### Hydrometeorological Data

Hydrometeorological data were sourced from the Unified COVID-19 dataset compiled by Badr and colleagues^16^, in which variables were in turn extracted from the second generation North American Land Data Assimilation System (NLDAS-2) and the fifth generation European Centre for Medium-Range Weather Forecasts (ECMWF) atmospheric reanalysis of the global climate (ERA5) at administrative unit centroids.^27–29^ Both datasets perform well in validation studies^28,30^ and are comparable to those used in retrospective infectious disease modeling^31^, with the advantage that their much shorter latency periods (4-6 days) make them better suited for prospective forecasting of disease dynamics.^16^ All available hourly, population-weighted ERA5 and NLDAS values since January 1, 2020 were extracted, aggregated to daily mean or total values, and matched by date and district to the R_*t*_ values. The following variables were included as the main exposures of interest based on their documented or hypothesized associations with SARS-CoV-2: near surface air temperature (°C)^32,33^; relative humidity (%)^34^; solar radiation (KJ/m^2^)^35^; total precipitation volume (mm)^36^; average 10-m above ground wind speed (m/s).^37^ In addition, average volumetric soil moisture (m^3^/m^3^) was included as a negative control exposure^38^, since it is a variable presumed to affect infectious disease transmission through its influence on pathogen survival on surfaces and fomites^31^, which is thought to be at most only a secondary mode of SARS-CoV-2 transmission.^39^ Specific humidity (kg/kg) estimates were excluded from the main analysis due to being highly correlated with temperature in this dataset (*ρ* = 0.88), and only included in a secondary analysis reported in the **supplementary appendix**.

### Covariate Data

The following variables (summarized in **Table 1**) were included as covariates to adjust for their potential confounding effects on the main associations of interest:

**Table 1:**
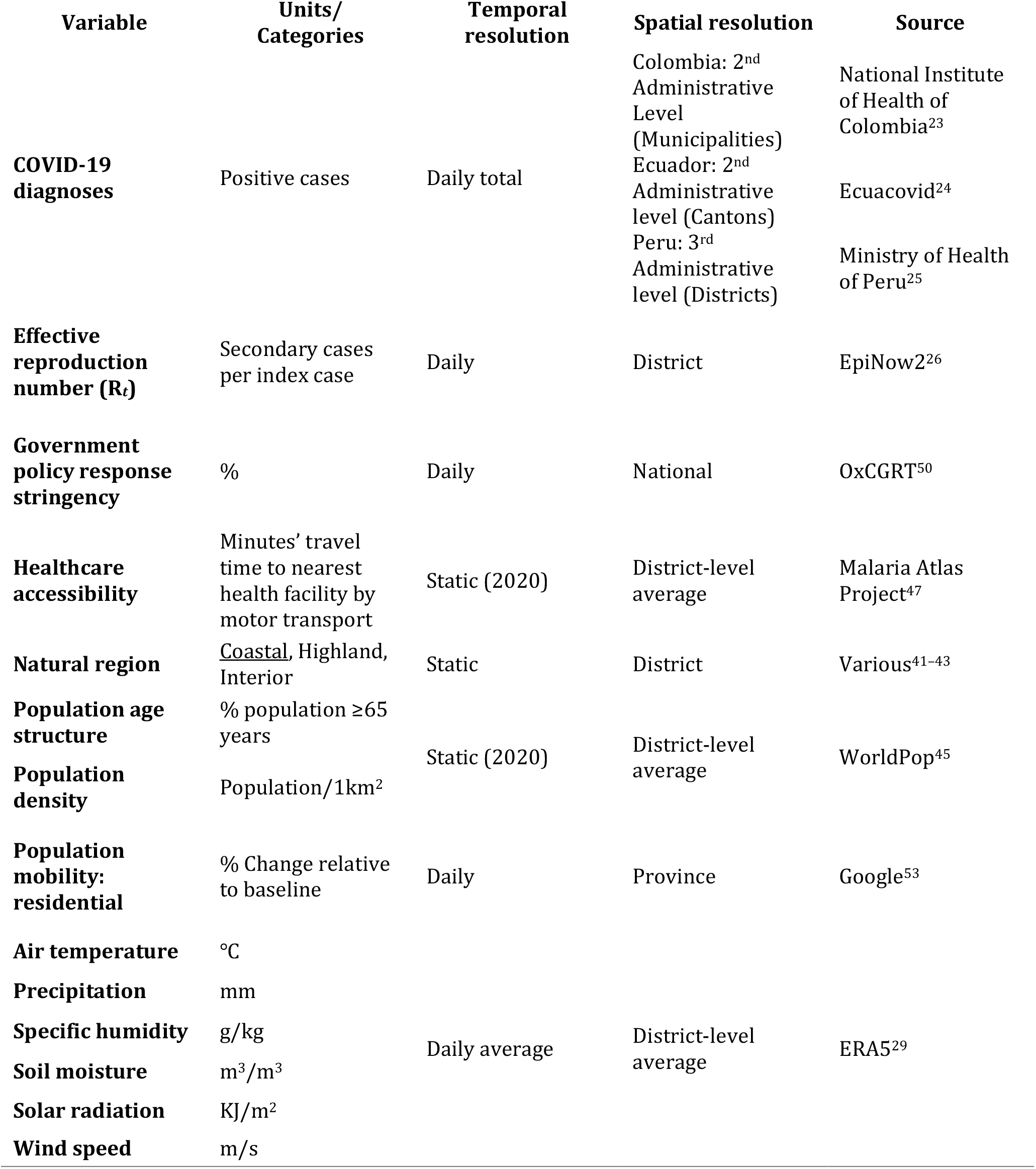
Definitions of variables used in the analysis.

| Table 1: Definitions of variables used in the analysis |  |  |  |  |
| --- | --- | --- | --- | --- |
| Variable | Units/<br>Categories | Temporal<br>resolution | Spatial resolution | Source |
| COVID-19<br>diagnoses | Positive cases | Daily total | Colombia: 2<sup>nd</sup><br>Administrative<br>Level<br>(Municipalities) | National Institute<br>of Health of<br>Colombia<sup>23</sup> |
|  |  |  | Ecuador: 2<sup>nd</sup><br>Administrative<br>level (Cantons) | Ecuacovid<sup>24</sup> |
|  |  |  | Peru: 3<sup>rd</sup><br>Administrative<br>level (Districts) | Ministry of Health<br>of Peru<sup>25</sup> |
| Effective<br>reproduction<br>number ($R_t$) | Secondary cases<br>per index case | Daily | District | EpiNow2<sup>26</sup> |
| Government<br>policy response<br>stringency | % | Daily | National | OxCGRT<sup>50</sup> |
| Healthcare<br>accessibility | Minutes' travel<br>time to nearest<br>health facility by<br>motor transport | Static (2020) | District-level<br>average | Malaria Atlas<br>Project<sup>47</sup> |
| Natural region | <u>Coastal</u>, Highland,<br>Interior | Static | District | Various<sup>41-43</sup> |
| Population age<br>structure | % population $\geq 65$<br>years | Static (2020) | District-level<br>average | WorldPop<sup>45</sup> |
| Population<br>density | Population/1km<sup>2</sup> |  |  |  |
| Population<br>mobility:<br>residential | % Change relative<br>to baseline | Daily | Province | Google<sup>53</sup> |
| Air temperature | °C | Daily average | District-level<br>average | ERA5<sup>29</sup> |
| Precipitation | mm |  |  |  |
| Specific humidity | g/kg |  |  |  |
| Soil moisture | m<sup>3</sup>/m<sup>3</sup> |  |  |  |
| Solar radiation | KJ/m<sup>2</sup> |  |  |  |
| Wind speed | m/s |  |  |  |

#### Natural regions

To account for potential residual confounding due to geographical and topographical differences across the three countries that may affect disease transmission^40^, their territories were grouped into three broad, cross-cutting ecological zones, based on the “natural region” categories used by the Peruvian national statistical authority - coastal, highland (the Andes), and interior (the Amazon and Orinoco basins).^41–43^ These were deemed to be less arbitrary from the point of view of transmission and meteorological dynamics than alternative groupings based on political divisions such as higher-level administrative units. The three regions are shown in **figure 3d**.

#### Population density

Densely populated urban areas are often struck earlier and harder by epidemics due to their roles as transport hubs and increased contact rates between susceptible and infectious individuals.^44^ Since sparsely populated areas may also differ systematically in the climate conditions that they experience, population density was included as a potentially confounding covariate in this analysis and calculated as the district-level zonal mean value extracted from the WorldPop raster of global population distribution.^45^

#### Population age structure

Because the symptomaticity and severity of SARS-CoV-2 infection increases with age^46^, areas with a larger proportion of their population in the more susceptible elderly age groups may have higher rates of case reporting and infectiousness. Population age structure varies geographically to a considerable degree, therefore the proportion of a district’s population that is over the age of 65 years was calculated from the WorldPop rasters of population per 5-year age group and included in the model.

#### Access to healthcare facilities

The time it takes to travel to a health facility to seek care also varies geographically as a function of population density, transport infrastructure and local topography.^47^ Connectivity has been shown to influence variation in SARS-CoV-2 outbreaks in sub-Saharan Africa^48^ and travel time to care-seeking might affect contact rates between infected and susceptible individuals or the probability that infected persons are treated and registered in health information systems. The district-level mean travel times to the nearest healthcare facility using motorized transport in 2020 were extracted using zonal statistics from the geographical estimates published by Weiss and colleagues.^47^

#### Government policy response data

The timing and stringency with which national governments introduced public health interventions such as travel restrictions, school closures and bans on gatherings and public events are major factors influencing geographical variation in the trajectory of the pandemic.^19,44,49^ The Oxford Covid-19 Government Response Tracker (OxCGRT) project collates information on numerous government policy responses into a publicly available database, assigns them scores reflecting their strictness and aggregates these into policy metrics including the stringency index^50^, which was included as a national-level, time-varying covariate in this analysis.

#### Population mobility

Compliance with NPI mandates and recommendations differ between subnational populations leading to variation in transmission risk.^51,52^ As a proxy indicator for compliance with social distancing, lockdown measures and travel restrictions, population mobility metrics were sourced from Google’s Community Mobility Reports.^53^ These indicators track trends in Android smartphone users’ movements over time relative to a pre-pandemic baseline, by subnational region and for six categories of location.^53^ The “residential” metric was used and merged with the database by date at the 1^st^ administrative unit level (hereafter generically referred to as “provinces”), since coverage was more complete at that level than for districts. This variable can be interpreted as the percent change in time spent in residential areas compared to before the pandemic, with a higher value therefore corresponding to greater population compliance with social distancing or lockdowns. The Google mobility dataset includes intentional gaps for unit-dates that don’t meet a quality and privacy threshold, which are to be considered “true unknowns”, so these intermittent missing values were substituted using linear interpolation by date within each province.^53^

### Statistical Analysis

Variables were merged based on district/province ID and date and highly skewed variables were normalized using Ordered Quantile (ORQ) transformation. A generalized additive mixed effects model (GAMM) was fitted to the R_*t*_ outcome assuming a Gaussian distribution, log link, and REML smoothing parameter estimation method. Cubic spline terms with 3 degrees of freedom were specified for all continuous exposure variables to account for non-linearity. Natural regions were modeled as a factor variable with the coastal region as the reference category. District-level random effects were specified to account for within-unit non-independence of the observations. The modeled, adjusted associations were visualized in partial dependence plots of R_*t*_ predictions across the range of values of each continuous exposure. Variable importance was assessed and ranked by computing the mean absolute accumulated local effects (ALE) of each predictor. To assess and compare relative effects, highly ALE-ranked variables were dichotomized at specific thresholds and otherwise identical GAMMs refitted to calculate the percent difference in R_*t*_ on unit-days above compared to below those thresholds.^54^ To quantify the variance explained by the hydrometeorological relative to the other variables, the R^2^ of the final model was compared to that of an otherwise identical model that included only the non-hydrometeorological predictors. Data processing, visualization, and analysis were carried out using R 4.0.3^55^, Stata 16^56^, and ArcMap 10.8.^57^

## Results

Data from the 3,212 mainland districts of the three countries were included for the 245-day period from May 1^st^ to December 31^st^, 2020, resulting in a dataset with a total of 786,940 unit-day observations. 564,738 (71.8%) observations were excluded due to having both zero cases and an estimated R_*t*_ value of between 0.95 and 1.05. A further 6,952 (3.6% of the remaining observations had missing mobility index data, leaving 184,870 complete observations to which the model was fitted. **Figure 1** shows choropleth maps of the geographical distribution of cumulative COVID-19 cases and average SARS-CoV-2 R_*t*_ (after applying exclusion criteria) summarized from daily values over the period of analysis. Neither showed a marked geographical pattern, though cumulative case burden (**figure 1a**.) exhibited notably lower values in the Peruvian highland districts, while many of the highest average R_*t*_ values (**figure 1b**.) were seen in the Ecuadorian and Colombian highlands. In Peru, the districts reporting more than 10,000 cases over the analysis period were in the Greater Lima Region as well as the other coastal cities of Trujillo and Chiclayo, while in Ecuador, aside from cantons of the three major cities of Guayaquil, Quito and Cuenca, the much less populous canton of Cañar also exceeded this threshold. Colombia experienced more numerous pockets of high cumulative cases in the major metropolitan municipalities of its highlands – Bogotá, Medellín, Cali – and Caribbean coast – Barranquilla, Cartagena – as well as several relatively smaller cities including Valledupar, Manizales, and Soledad.

**Figure 1:**
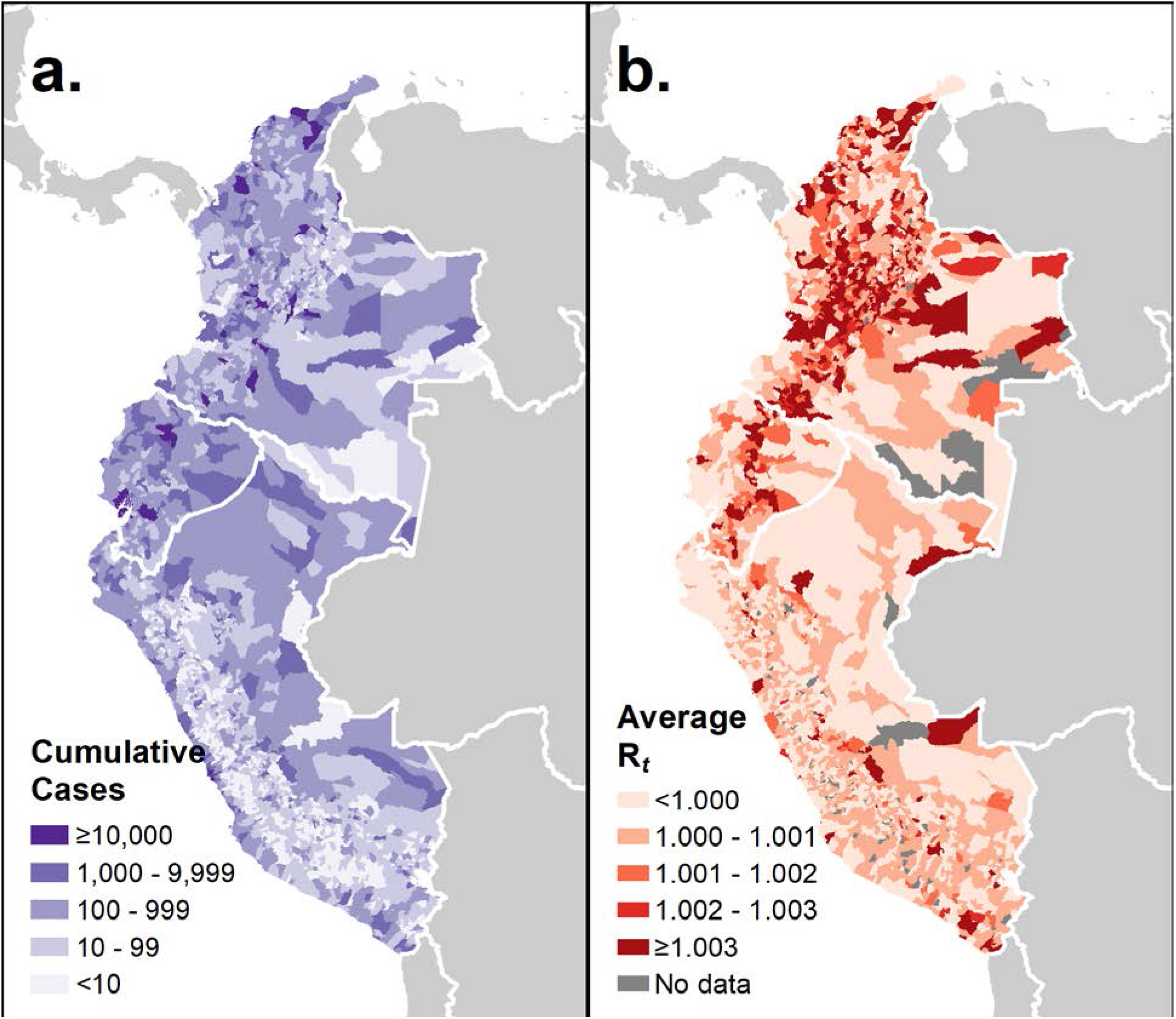
District-level geographical distribution of cumulative reported COVID-19 cases and estimated SARS-CoV-2 reproduction number (R_*t*_) in Colombia, Ecuador, and Peru (May 1^st^ – December 31^st^, 2020).

**Figure 2** shows equivalent choropleth maps for averages of the six hydrometeorological variables. The lowest average temperature values (**figure 2a**.) occurred along the Andes, particularly in the southern Peruvian stretch, while the highest occurred in the low-lying interior regions of the Amazon and Orinoco basins, as well as coastal Ecuador and Colombia. relative humidity (**figure 2b**.) and soil moisture (**figure 2c**.) exhibited similar spatial distributions with the highest average values in the interior areas of the three countries and along the Colombian coast, except for the arid Guajira peninsula, which had very low soil moisture content of <0.2 m^3^/m^3^. Other areas of very low humidity and soil moisture included Peru’s coastal Sechura Desert and Colombia’s central Tatacoa Desert, as well as small pockets along the Ecuadorian coast. Average wind speeds (**figure 2d**.) exceeded 1 m/s along most of the Pacific and Caribbean coasts, and the high elevation Andean districts, while the mid-elevation windward and leeward Andean districts tended to have wind speeds of less than 0.5 m/s as did parts of north central Colombia. Precipitation distribution (**figure 2e**.) largely mirrored that of soil moisture and relative humidity with the Guajira, Sechura and Tatocoa Deserts experiencing low average daily rainfall of <1.5mm, and the Pacific coast and interior of Colombia exhibiting the wettest conditions. A belt of high solar radiation (**figure 2f**.) extended along Peru’s coast, which widened in the southeast to incorporate highland areas of the Andean Plateau. The only other area of comparable radiation levels was on Colombia’s Guajira peninsula.

**Figure 2:**
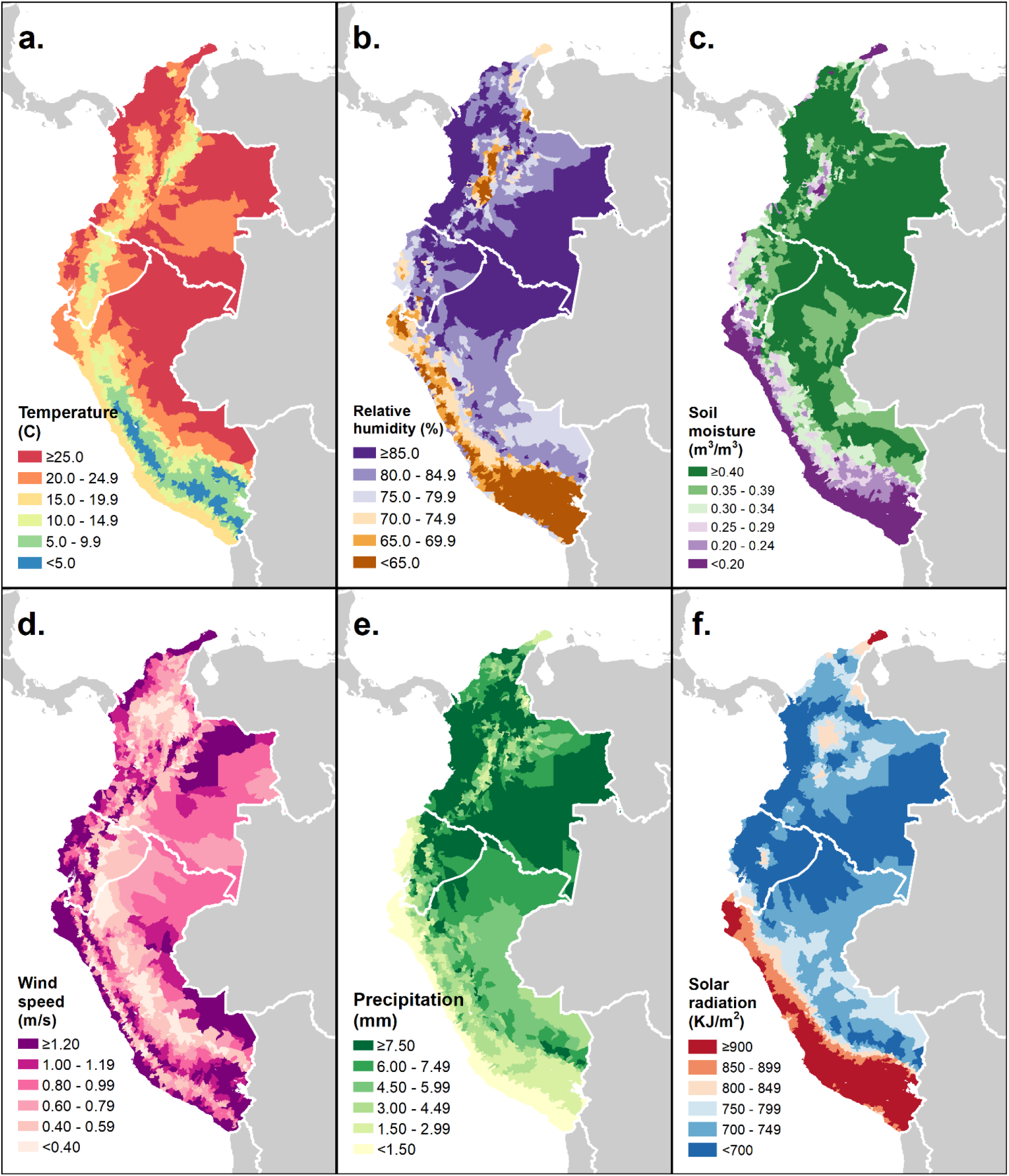
District-level geographical distribution of six hydrometeorological variables in Colombia, Ecuador, and Peru (mean of daily averages May 1^st^ – December 31^st^, 2020).

**Figure 3:**
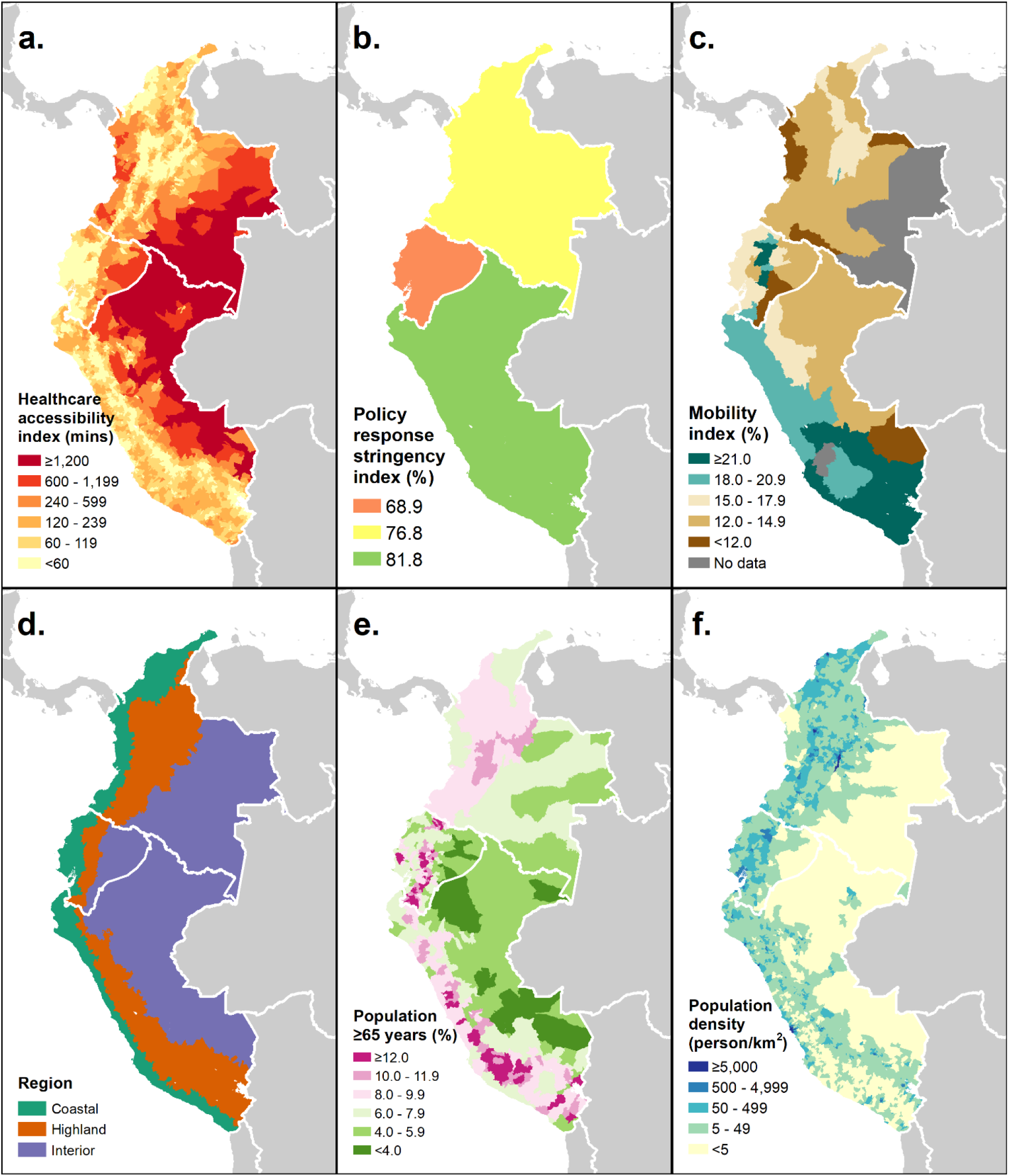
District-level geographical distribution of 6 covariate variables in Colombia, Ecuador, and Peru (May 1^st^ – December 31^st^, 2020).

**Figure 3** shows equivalent maps for the non-hydrometeorological covariates, including the extents of the three natural regions (**figure 3d**.). The population (**figure 3f**.) of the three countries is concentrated along the coasts and highlands, with the exceptions of Colombia’s sparsely populated Darién Gap and the high elevation districts of Peru’s southeast Andes. The population of the countries’ interiors is not only sparse, but also most highly skewed towards the younger age groups (**figure 3e**.), while the highlands and Peru’s coastal plains have some of the districts with the highest percentage of the population over 65 years. Access to healthcare, as measured by average travel time to the nearest health facility by motor transport (**figure 3a**.), tends to vary inversely with population density, with the lowest levels of accessibility seen in the interior regions and along Colombia’s Pacific coast, the coasts and Andean highlands having the majority of districts with an average travel time of under an hour. Over the period from May to December 2020, Peru had the most, and Ecuador the least stringent policy response to the pandemic on average (**figure 3b**.). Average time spent in residential locations (**figure 3c**.) was highest (meaning mobility was lowest) in Peru’s coastal and southern highland areas, in Ecuador’s central highlands and in the Capital District of Bogotá, Colombia and lowest in provinces along the countries’ land borders and Colombia’s Pacific coast.

**Figure 4** visualizes the adjusted associations from the GAMM. Precipitation and wind speed were ORQ-transformed due to their skewed distributions. All six variables were highly statistically significantly associated with the outcome at the *α*<0.0001 level. Temperature’s effect (**Figure 4a**.) on district-level R_*t*_ was negligible in size, taking on a slight sinusoidal shape across the range of the variable’s distribution. Precipitation (**Figure 4b**.) showed a broadly lop-sided U-shaped association with R_*t*_ with the lowest predicted value in the mid-range and the highest at the upper extreme. The effect of soil moisture (**Figure 4c**.) was direct below a moisture value of approximately 0.25 m^3^/m^3^, forming a plateau above that threshold. Solar radiation’s association (**Figure 4d**.) took the form of a descending arc with the inverse relationship most marked above a threshold of approximately 700 KJ/m^2^, and the lowest predicted R_*t*_ for any of the hydrometeorological variables (R_*t*_ <0.97) occurring at the upper radiation extreme of close to 1,500 KJ/m^2^. relative humidity (**Figure 4e**.) had the largest magnitude effect size of any hydrometeorological variable, with increasing humidity mostly predicting decreasing R_*t*_ (except for a plateau from around 70 – 80% relative humidity) and a difference in predicted R_*t*_ of approximately 0.05 between the extremes of the distribution. The association of wind speed with R_*t*_ (**Figure 4f**.) was small in magnitude and inverse in approximately the upper tercile of the ORQ-transformed distribution.

**Figure 4:**
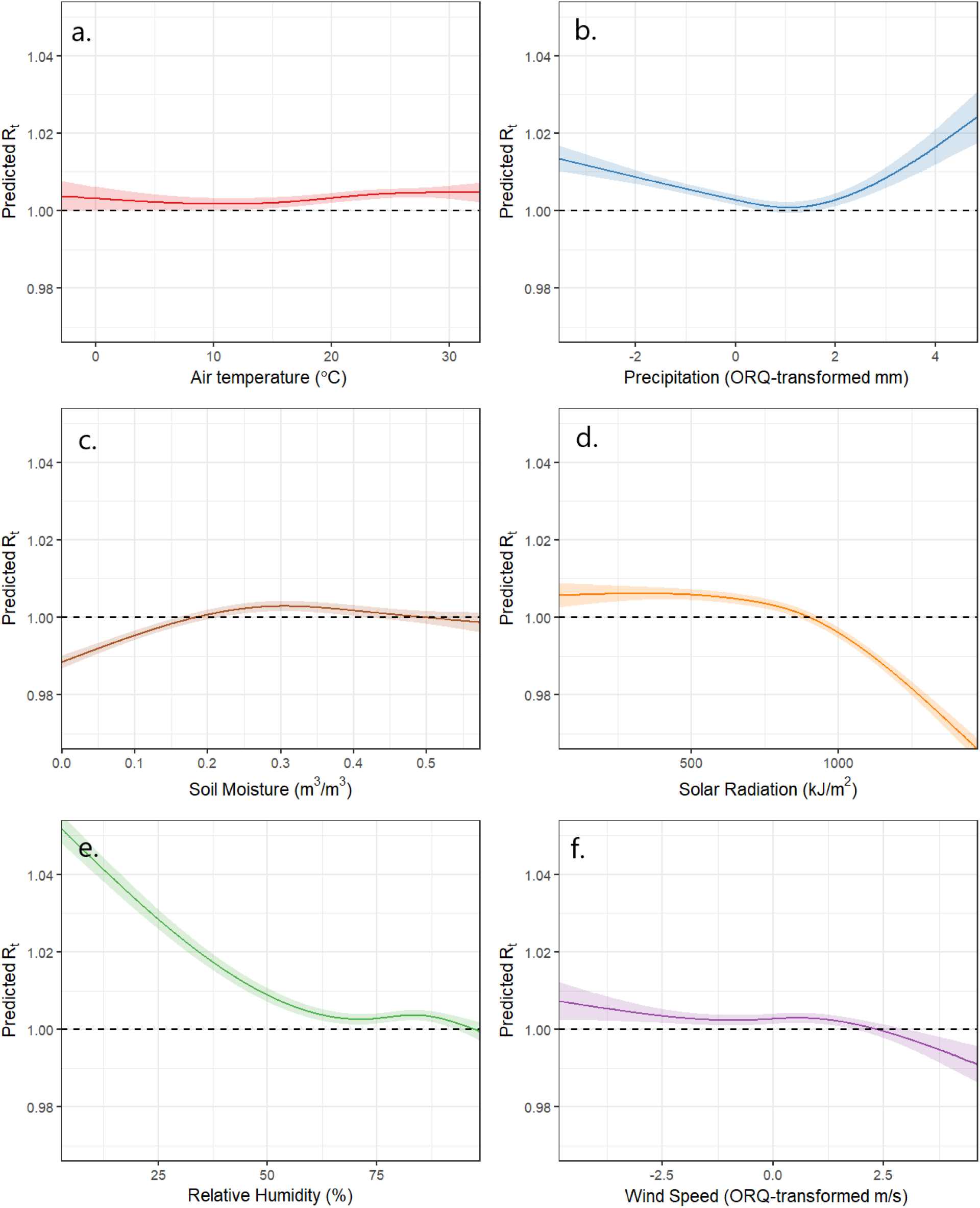
Adjusted associations between 6 hydrometeorological variables and daily COVID-19 reproduction numbers R_*t*_ values predicted by generalized additive mixed effects model.

**Figure 5** shows the equivalent associations for the five continuous, non-hydrometeorological covariates and the coefficient estimates for the two comparison natural region categories relative to the “Coastal” reference category. Population density and health facility accessibility were ORQ-transformed. R_*t*_ increased with longer travel times to health facilities (**Figure 5a**.) from a value of 0.98 at the shortest time to just above 1 in the upper half of the ORQ-transformed accessibility distribution. The government response index (**Figure 5b**.) had a negligible effect on SARS-CoV-2 R_*t*_ and did not predict a value below 1 at any value. Population mobility (**Figure 5c**.) had a large, steep inverse association with population mobility below a percent change of 10% -meaning that R_*t*_ increased when time spent in residences decreased little or increased relative to the pre-pandemic baseline - and a steady direct relationship above that threshold. The adjusted effects of the categorical natural region variable (**Figure 5d**.) were small and non-significant. Population density (**Figure 5e**.) had a direct association with the outcome with the most densely populated districts having an adjusted predicted R_*t*_ of >1.05, along with population mobility, one of the largest effect sizes observed in this analysis. Though statistically significant at the *α*<0.001 level, the effect of population age structure (**Figure 5f**.) was negligible consisting of a shallow, direct association within a short range of values between roughly 6% and 11% of the population aged over 65 years.

**Figure 5:**
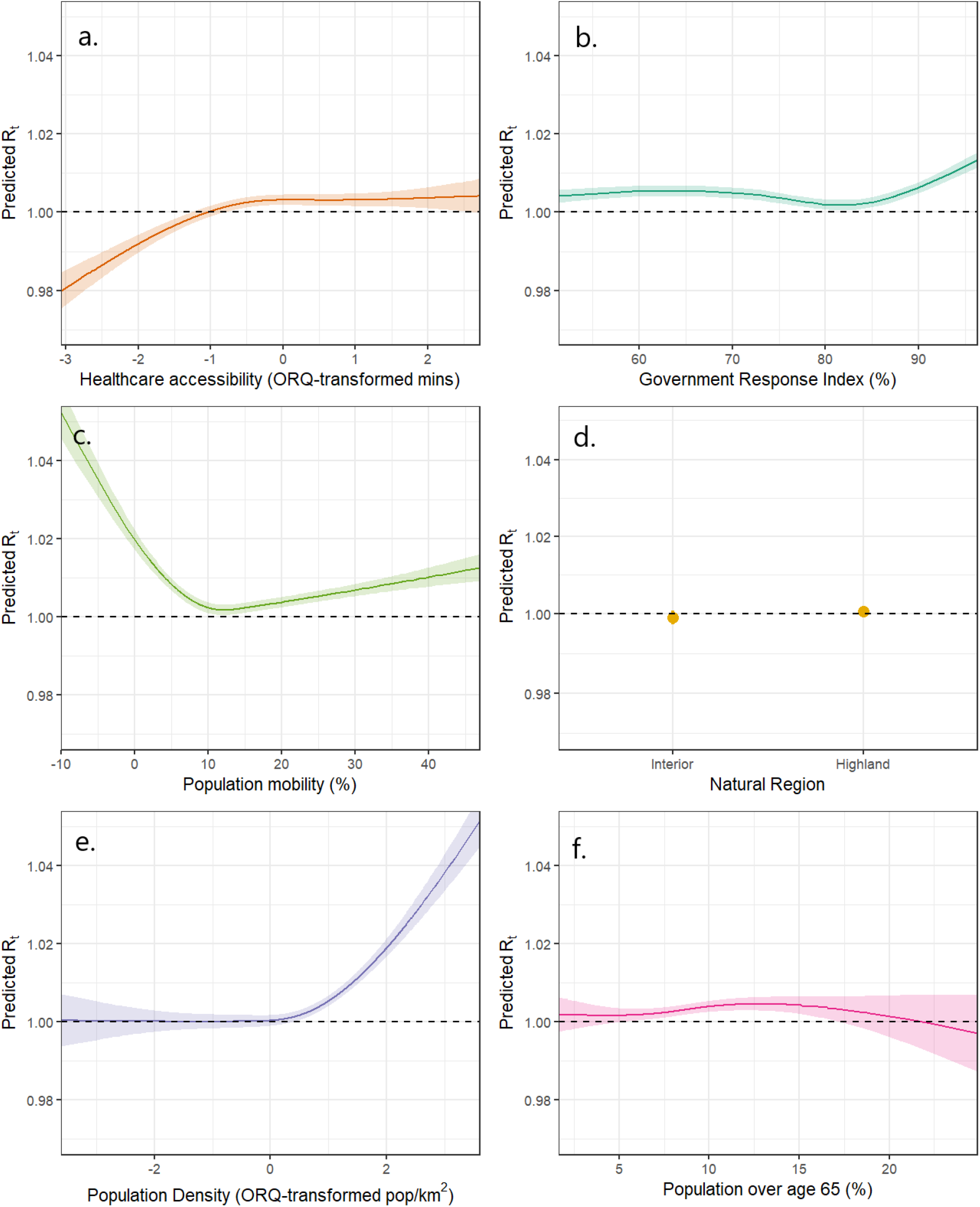
Adjusted associations between 6 covariate variables and daily COVID-19 reproduction numbers R_*t*_ values predicted by generalized additive mixed effects model.

The final model explained 3.9% of the variance in the daily district SARS-CoV-2 R_*t*_, compared to 2.6% by an equivalent model that excluded the hydrometeorological variables. ALEs for all variables were correspondingly small (**supplementary table S1**), with population density ranking highest in terms of contribution to R_*t*_ (ALE = 0.007) followed by solar radiation (ALE = 0.004), the highest ranking of the hydrometeorological variables. In models in which the highest ALE-ranked variables were dichotomized (**table S1**), differences in average predicted R_*t*_ for unit-dates on either side of variable-specific thresholds were also modest. Average adjusted R_*t*_ was 1% lower on days in which relative humidity was higher than 50%, compared to less humid days, but 1% higher when soil moisture was above 0.1 m^3^/m^3^. Days in which solar radiation exceeded 1,000 KJ/m^2^ had 1.3% lower R_*t*_, while the equivalent differences for districts in which average travel time to health facilities was more than half an hour and with population density of more than 100 pop/1km^2^ were, respectively, a 0.2% and a 0.4% increase, and on days in which mobility was reduced by 10% or more relative to pre-pandemic levels, R_*t*_ fell by an average of 0.3%

## Discussion

The impacts of the COVID-19 pandemic on families, societies and institutions have been incalculable. Furthermore, the notable spatiotemporal variability in these impacts are seemingly not fully attributable to population susceptibility and health system factors alone^58^, implicating a potential influence of climate and environment on the transmission and survivability of SARS-CoV-2.^12^ Early surveys of the evidence base highlighted a paucity of findings from the Global South and tropical regions, insufficient spatiotemporal scope and resolution in analyses, and a failure to account for confounding from non-climatological factors.^12,20,22,59^ More recently, as attempts to track the pandemic have coalesced into a wide variety of open datasets and online interfaces^15,16,60^, researchers have begun to address these knowledge gaps. Numerous recent studies have assessed effects on COVID-19 outcomes adjusting for multiple hydrometeorological variables^35,61^ and other covariates including population density^44^, age structure^62^, NPI compliance^33,63^ and government interventions^64^, while others have focused on single countries in equatorial regions^65,66^ or multiple countries and locations spanning wide latitudes and both hemispheres.^19,67,68^ This study is the first to bring together all these elements and at a high temporal resolution, multiple, cross-cutting spatial scales and for three neighboring countries that, despite including diverse populations and ecologies, share important commonalities in their pandemic experiences.

The ancestor of the SARS-CoV-2 index virus likely evolved through transmission among bats living in cool, dark, crowded caves.^69,70^ The primary direct, person-to-person mode of transmission of the pathogen is via virus-laden aerosols exhaled by infectious individuals, while an indirect route via contact with contaminated fomites is thought to make a minor contribution.^39,71^ Small-scale atmospheric conditions such as the temperature, pressure and humidity of the air affect the rates at which aerosolized respiratory droplets are formed, suspended, and dispersed and thus influence disease transmission in complex ways.^31,34^ The negative association of relative humidity on SARS-CoV-2 R_*t*_ identified here, among the largest absolute effect sizes of the hydrometeorological variables analyzed (though lower ranking by ALE), is consistent with one of the most widely documented of the disease’s environmental sensitivities as well as current understanding regarding the virus’ modes of transmission.^34,72^ Whether quantified by absolute or relative measures, humidity has been shown to be an influential COVID-19 driver across many contexts^37,73^, with very dry atmospheric conditions appearing to favor transmission as has been shown for other respiratory^74^ and non-respiratory^54^ viruses. When expelled into dry air, respiratory microdroplets quickly shrink due to evaporation of their liquid content, allowing them to suspend aloft for longer and increasing their viral particle concentration.^72,75^ relative humidity also has a separate U-shaped association with SARS-CoV-2 viability outside the human host, with its lowest viability occurring around 60% air saturation and its highest at the extremes.^34^ Competing effects of decreasing transmissibility and increasing viability in the upper humidity extreme are consistent with the plateau effect seen in these results at relative humidity >70%.

Though there is less consensus surrounding the effect of temperature, it is widely supposed to have an association similar to that of humidity, and indeed numerous studies have reported decreasing COVID-19 risk with increasing temperatures.^61,62,73^ While this might at first glance seem to be at odds with the negligible and non-linear effect found in this analysis, comparisons with results specifically from other tropical settings suggest a more nuanced picture. One such study within a single season in Singapore^66^ (January to April 2020) found a strong and significant direct association between temperature and COVID-19 case numbers, while another, also of a tropical, equatorial South American country (Brazil), found opposing effects of temperature in the March to May period (direct) compared to from June to August (inverse).^65^ Another study of >400 cities across a wider range of latitudes in both the northern and southern hemispheres found a more complex, sinusoidally shaped relationship of temperature to transmission.^19^ Since the domain of this analysis spanned only tropical latitudes either side of the equator, the null-like finding for temperature could plausibly be the result of competing effects between the two hemispheres at different times over the year canceling each other out. However, marginal temperature effects predicted by multivariable models may be sensitive to the choice of humidity metric. The 400 city study adjusted for both relative humidity and absolute humidity^19^, while a study of US counties that adjusted temperature for specific humidity found still another complex and non-linear effect shape.^18^ Supplementary **figure S1** compares the results of this model with an otherwise identical one that substituted specific humidity for relative humidity, and reveals a somewhat larger magnitude and direct effect of temperature in the specific humidity model. Temperature and specific humidity are closely related variables and exhibited multicollinearity in this dataset (a variance inflation for specific humidity of >10 in models that include temperature). Moreover, certain combinations of their values (e.g., low temperature with high specific humidity) simply do not occur naturally, so attempts to visualize effects of variations in one parameter while holding the other constant at its mean value are in some senses abstractions.

This analysis also identified a sizeable, inverse association of solar radiation on COVID-19 transmission consistent with numerous other studies^37,44,68^, most notably by Ma and colleagues, who also found this to be most pronounced above a threshold of ∼1,000 KJ/m^2^.^18^ Such findings have been interpreted as reflecting the deactivating effect of sunlight on SARS-CoV-2 virions as has been observed in laboratory conditions either in aerosols^76^, on surfaces^77,78^ or in mucus.^79^ However, commentators have noted the difficulty of disentangling a direct effect of sunlight on the disease agent itself, from its confounding effect on host behaviors, such as rainy or cloudy weather driving people to congregate indoors, thereby increasing contact rates.^54,68^ The fact that this effect was observed with adjustment for precipitation lends credibility to the supposed direct effect. Similarly, a substantial effect of precipitation was observed with adjustment for population mobility, the highest predicted R_*t*_ occurring at the high end of the precipitation distribution, the lowest in the mid-range and a secondary peak on rainless days. Given the absence of a waterborne route of transmission, it is tempting to attribute this to residual, unobserved confounding from host behaviors that are incompletely captured by the mobility variable, rather than a direct, causal impact of rainfall on virus dispersal. However, the role of aerosolized particles from wastewater cannot be ruled out.^80^ In Andean countries like these with wide inequities in sanitation coverage, many community-level environments are characterized by poor sewerage infrastructure^81^, where open wastewater canals serve the dual functions of drainage for rainwater runoff, and conveyance of effluent discharge from household latrines.^82^ Such basic systems are easily overwhelmed by heavy rain events^83^, which may promote the creation of airborne contaminated bioaerosols in which infectious pathogens can remain viable, as has recently been demonstrated for several enteropathogens including viruses.^84^

Soil moisture was included as a negative control exposure yet its observed effect on R_*t*_, though small in absolute terms, was larger than that of government policy, population age structure and natural region. A possible explanation is that soil moisture serves as a proxy for the general moisture retention of all surfaces, and that virus particles expelled in aerosolized droplets may remain viable for longer if they settle on a surface that permits them to retain their surrounding moisture.^85^ A small, inverse effect of wind speed above a threshold high in the distribution was identified, consistent with that identified by Clouston and colleagues.^86^ While several other studies have also found inverse effects^61,63^, and still others have reported direct^37,64,67^, or negligible^65^ effects of wind speed, it seems highly plausible that faster wind speeds suppress transmission of SARS-CoV-2 in outdoor environments by increasing air circulation and dispersing infective aerosols away from susceptible individuals, much as ventilation does in indoor environments.^80,86^

The modeled effects of several non-hydrometeorological variables were consistent with the a priori hypotheses justifying their inclusion. Transmission was highest in densely populated districts, presumably due to higher contact rates, and lowest in districts with shorter travel time to health facilities, perhaps due to improved access to diagnosis and case management shortening the period between disease onset and isolation, or to unresolved confounding by latent urban status. On days in which time spent in residences was at least 10% more than was typical before the pandemic (a proxy for lockdown compliance), R_*t*_ was statistically significantly reduced, though by less than 1% and not to a level below 1, that if sustained would eventually bring transmission under control. The proportion of the population that was elderly had no substantial impact on R_*t*_, likely because old age is less a risk factor for infectiousness or susceptibility to infection but rather for more severe COVID-19 outcomes once infected. It is striking that greater government response stringency had no effect on reducing R_*t*_ and even appeared to increase it slightly in the upper extreme. This may reflect that government response is often slow and largely reactive to surges in cases, or that it has little impact over and above that which is mediated by individual behavior change and compliance, factors captured by the mobility variable.

Besides those inherent to unit-level, ecological studies, this analysis is subject to further limitations. The high level of geographical disaggregation meant that there was an inflated number of unit-dates on which zero cases were reported or that had uninterpretable R_*t*_ values. This meant that the distribution of the outcome values was narrower and the effect sizes smaller than those reported in other comparable analyses, some by almost an order of magnitude.^19,35^ However, given that environmental conditions vary on a very small scale and that case data were available at such high resolution, this was deemed a justifiable tradeoff. The restriction of the analysis to the eight-month May to December period meant that it did not capture a full annual cycle of either weather conditions, or virus circulation, but was necessary to avoid having to account for vaccine and variant introduction.

In conclusion, COVID-19 transmission is sensitive to spatiotemporally varying hydrometeorological conditions in these three countries of tropical Andean South America, even after adjusting for other potential confounders including both static and time-varying variables, and at multiple cross-cutting scales. Dry atmospheric conditions of low humidity increased, and higher solar radiation decreased district-level SARS-CoV-2 reproduction numbers. While several commentators have cautioned that the effects on transmission of climatological conditions are likely to be modest compared to factors such as NPI compliance,^59,68^ these findings in fact show their influence to be of a comparable magnitude in several cases and even greater than that of government response and population age structure. However, in absolute terms these effects, though significant, are modest and do not explain the excess disease burden experienced in some parts of this region during the first wave of the pandemic. As SARS-CoV-2 settles into indefinite endemic circulation, it may be feasible to incorporate weather monitoring into disease surveillance and early warning systems alongside other more costly activities such as wastewater or population seroprevalence surveillance for anticipating case surges and allocating resources. Furthermore, population health interventions that encourage the public to exercise greater precautions on cloudy or dry days could also be considered. However, the high proportion of variance in COVID-19 transmission that remains unexplained even after accounting for population factors and NPIs (>96%) are striking, as are the negligible relative effect sizes of <2%, which are far surpassed by those of interventions such as vaccination (53 – 94%^87^) and mask wearing (19%^88^), all of which should serve as cause for caution when attempting to predict near-term changes in transmission risk.

## Data Availability

The data and R code used in this analysis are provided in this GitHub repository https://github.com/joshcolston/Colston_COVID-19_in_the_Andes.

## Acknowledgements

The research presented in this article was supported financially by a COVID-19 supplement to NASA’s Group on Earth Observations Work Programme (16-GEO16-0047) to Drs. Zaitchik and Kosek. Additional funding was obtained from the Centers for Disease Control and Prevention (U01GH002270) to Dr. Kosek. The funders played no role in study design, the collection, analysis, and interpretation of data, the writing of the report, or the decision to submit the paper for publication. The findings and conclusions of this report are those of the authors and do not necessarily represent the official position of the funders. We are grateful to Ecuacovid, a project that provides a set of raw data extracted from reports on the national COVID-19 situation from Ecuadorian health authorities.

## Author contributions

JMC, MNK and BFZ conceived of the study. MNK and BFZ secured funding for the research. JMC, PH, NHN, YTC, HB, and DNM carried out data processing, analysis, and visualization. MNK and AQ provided initial data. GHK, LMG, AQ and FSS provided interpretation and writing support.

