## Supplementary material for "Effects of hydrometeorological and other factors on SARS-CoV-2 reproduction number in three contiguous countries of Tropical Andean South America: a spatiotemporally disaggregated time series analysis"

### Supplementary materials

**Figure S1:** Comparison of adjusted associations between temperature and humidity and daily COVID-19 reproduction numbers $R_t$ values predicted by the main model, which used relative humidity (a. and b.), an otherwise identical one that substituted it for specific humidity (c. and d.).

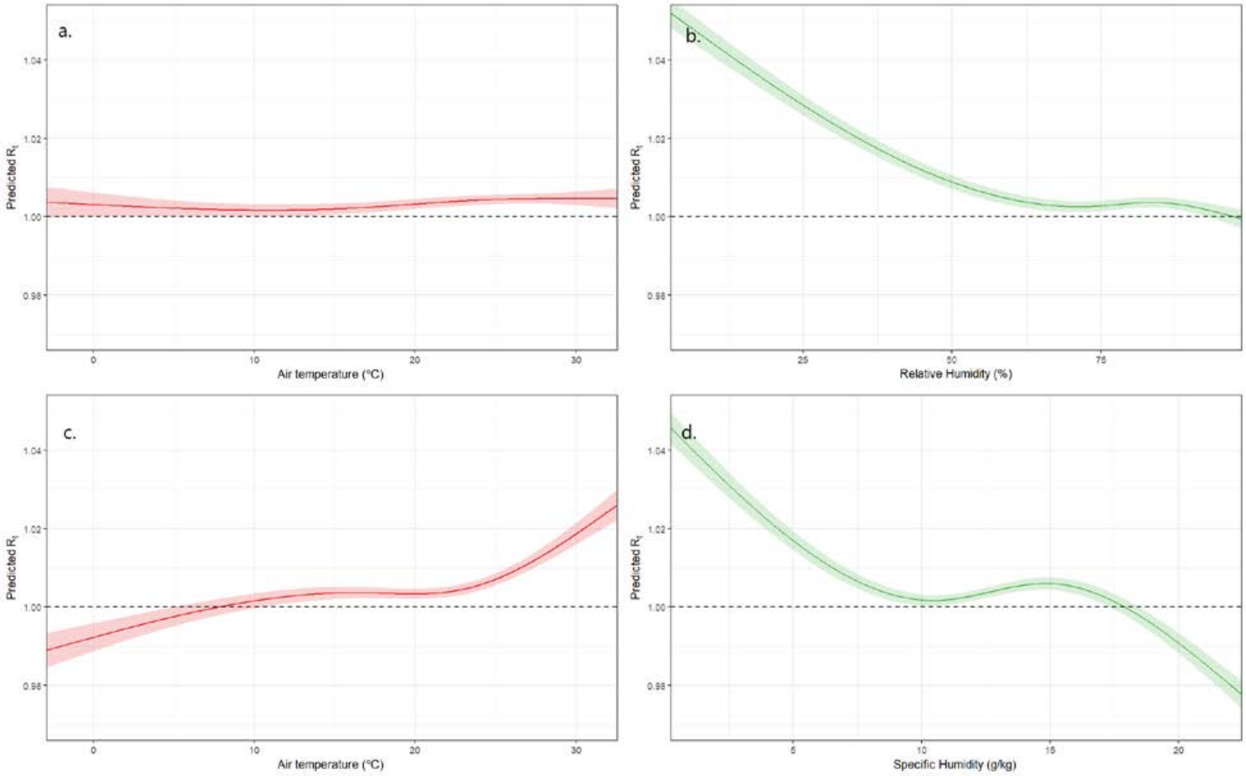

**Table S1: Accumulated local effects (ALE) of the 6 highest-ranked variables and predicted values of and percent difference in SARS-CoV-2 $R_t$ estimated from GAMMs in which they were dichotomized at variable-specific thresholds but were otherwise fitted identically to the main model.**

| Variable | ALE | Threshold | % Above threshold | Predicted $R_t$ below threshold | Predicted $R_t$ above threshold | % $R_t$ Difference |
|---|---|---|---|---|---|---|
| **Population Density** | 0.007 | 100 pop/1km$^2$ | 18.5 | 1.000 (0.999, 1.001) | 1.004*** (1.003, 1.006) | 0.4 |
| **Solar Radiation** | 0.004 | 1,000 KJ/m2 | 12.8 | 1.005*** (1.004, 1.006) | 0.992*** (0.990, 0.993) | -1.3 |
| **Healthcare Accessibility** | 0.004 | 30 mins | 76.2 | 1.001** (1.001, 1.003) | 1.004*** (1.002, 1.006) | 0.2 |
| **Population Mobility** | 0.004 | 10% change | 89.3 | 1.006*** (1.005, 1.007) | 1.003*** (1.001, 1.005) | -0.3 |
| **Soil moisture** | 0.003 | 0.1 m$^3$/m$^3$ | 88.4 | 0.995*** (0.994, 0.996) | 1.010*** (1.009, 1.011) | 1.0 |
| **Relative Humidity** | 0.003 | 50% | 91.3 | 1.011*** (1.010, 1.013) | 1.001*** (0.999, 1.004) | -1.0 |

*** p<0.001, ** p = 0.001 – 0.01, * p = 0.01 – 0.05

The full dataset and code for fitting the main model are available at the following GitHub repository https://github.com/joshcolston/Colston_COVID-19_in_the_Andes
